# Edematous Type of Malnutrition Takes Longer Recovery Time Compared to Severe Wasting (Marasmus): Findings on 6-59 months old children treated for Severe Acute Malnutrition After the War in Northern Ethiopia, 2023: Prospective Longitudinal Study

**DOI:** 10.1101/2023.10.20.23297307

**Authors:** Wagnew Tesfay, Mebrahtu Abay

**Author notes:** Corresponding author: Aksum, Ethiopia.

## Abstract

**Background:** Severe acute malnutrition (SAM) is still having public health significance by attributing almost fifty percent of the estimated ten to eleven million deaths encountered in less than five-years old children, imposing nine-fold likelihood of death compared with well-nourished children of similar age group. It increases terrifyingly and become lethal during conflicts due to lack of food, compromised water supply and hygienic practices along with insufficient healthcare services.

**Methods:** Hospital-based prospective cohort study was conducted using regularly collected programme data of children admitted to the stabilization center in Suhul general hospital from January 1^st^, 2023 up to July 31^st^, 2023. To collect the data, pretested structured questionnaire was generated referring to the national SAM management protocol. Then collected data get coded and entered into Microsoft Excel spreadsheet 2016 version. All forms of analyses were done using statistical package for social sciences (SPSS) version 25.

**Results:** From the total 168 children aged 6-59 months enrolled in the study, ninety-four per cent of them were recovered and transferred to the outpatient therapeutic programme while the rest six per cent of the participants were censored. Appetite test (AHR = 1.874; 95% CI: 1.180-2.978), level of dehydration (AHR = 0.538; 95% CI: 0.361-0.800 for some/moderate dehydration and AHR = 0.250; 95% CI: 0.128-0.489 for severe dehydration or shock), diagnosis at admission (AHR = 0.452; 95% CI: 0.294-0.694), blood transfusion (AHR = 5.559; 95% CI: 2.419-12.773), type of antibiotics (AHR = 0.365; 95% CI: 0.192-0.692) and nasogastric tube feeding (AHR = 0.531; 95% CI: 0.372-0.758) were declared significant predictors of recovery time.

**Conclusion:** Bottom line of the study shows the inpatient therapeutic feeding center has met the agreed indicators for nutrition interventions during humanitarian crises. We recommend the hospital leadership, the regional health bureau and other humanitarian agencies to stress on training clinical workforce directly involved in patient management and care.

## Background

The World Health Organization (WHO) defines severe acute malnutrition (SAM) as having severe wasting (measured by mid-upper arm circumference (MUAC) of less than 11.5 centimetre in children 6 to 59 months old, and/or weight-for-length/height (WFL/H) below -3 Z scores in kids 0-59 months of age) and/or bilateral pitting oedema (1).

Acute undernutrition is affecting more than 45 million under-five years old children worldwide, of whom about 14 million are severely malnourished (2–4). Although significant and productive efforts made to tackle it, SAM is still having public health significance by attributing almost 50% of the estimated 10 to 11 million deaths encountered in less than five-years old children, imposing nine folds likelihood of death compared with well-nourished children of similar age group (5,6). SAM increases terrifyingly and become lethal during conflicts due to lack of food, compromised water supply and hygienic practices along with insufficient healthcare services (7). All population segments are affected by acute malnutrition during emergencies, but SAM it is main cause of morbidity and mortality in infants and young children (8).

The armed conflict between Tigray regional forces and the federal government of Ethiopia erupted in November the 4^th^, 2020 has led to mass displacements and destruction of healthcare system in the region. As a result, with six and twenty two per cent of SAM and moderate acute malnutrition (MAM) rates respectively, nutritional status of children under-five years of age reached above the emergency threshold level for acute malnutrition (9,10). The regional rate of SAM before the conflict was about one per cent.

Recent studies in Ethiopia showed that rates of recovery are below the standard set for stabilization center (SC) (11,12) while others concluded with findings in line with the standards (13–15). Similar study in Uganda also reported unacceptably high death rate (16). Age, breastfeeding, immunization and presence or absence of co-morbidities during admission were associated with recovery time and related outcomes of SC (11,14,15). Co-morbidities like diarrhea, pneumonia, human immune-deficiency virus (HIV) sero-status and anemia were found as independent predictors of SC treatment outcomes (17,18). Variables like type of SAM, intravenous (IV) fluid resuscitation, nasogastric (NG) tube feeding, appetite and admission status were also significantly associated with therapy outcomes in SC (13,19–21).

During conflicts, research plays decisive role in understanding the main challenges faced by humanitarian organizations and enables them make scientifically informed decisions (22,23). Prolonged stay in SC is associated with a higher risk of hospital acquired infection which can raise the increased risk of death as well. Meanwhile, as a quality service indicator, recovery time and other related outcomes of nutrition interventions during humanitarian situation should be monitored to get better results of efforts and to take further actions based on evidence. Nevertheless, very little is known about the SC performance indicators in comparison against the standards during humanitarian interventions. This study would be the first of its kind since the armed conflict in the region broke out. Therefore, aim of this study is to avail evidences about recovery time and other performance indicators in Suhul general hospital, Tigray region, Northern Ethiopia.

## Methods and materials

### Study design

Health institution based prospective cohort study was conducted using regularly collected programme data of children admitted to suhul general hospital SC.

### Context and period of study

Located 1087 kilometers north of Addis Ababa, Shire town is the capital of north west zone of Tigray regional state and main host for internally displaced persons (IDPs). The inpatient therapeutic feeding center (ITFC) in Shire’s Suhul hospital has been the main management center for children suffering from complicated severe undernutrition, both for host and IDPs residing in the zone. Like almost all healthcare facilities in the region, this hospital has experienced extensive looting of medical supplies and equipment, electrical installations and destruction of water supply system. The cessation of hostilities agreement between the warring bodies signed in Pretoria paved the way for humanitarian organizations to access the region as well as the zone. As a result, the paediatrics ward of the general hospital, including the ITFC, has been receiving medical supplies and equipment as well as technical support from United Nations’ agencies and other international non-governmental organizations (NGOs).

Admission, treatment and discharge procedures were done as per the Ethiopian federal ministry of health (FMoH) SAM management guideline (24). Pediatrician and general practitioners were doing ward rounds on a daily basis to diagnose, prescribe or revise medications and other interrelated decisions while the nurses and nutrition nurses were assigned to do appetite test and move children from Phase-I to transition phase and phase-II. Collective and individual infant and young child feeding (IYCF) counselling sessions were given for mothers or guardians about conditions of their child, sanitation and hygiene practices and the necessary follow up until discharged from the ward. This study was conducted from January 1st, 2023 to July 31, 2023.

### Source and Study population

#### Source population

All 6-59 months old children admitted to suhul general hospital SC, both from host and displaced communities.

#### Study population

All 6-59 months old children admitted to suhul general hospital SC, both from host and displaced communities from January 01, 2023 to 31^st^ of July, 2023.

#### Study unit

Severely undernourished 6-59 months old child hospitalized to the stated SC from January 01, 2023 to 31^st^ of July, 2023.

### Inclusion and exclusion criteria

#### Inclusion criteria

All 6-59 months old children hospitalized to the SC both from host and displaced communities from January 01, 2023 to July 31, 2023.

#### Exclusion criteria

Children less than six months old, those with ready-to-use therapeutic food (RUTF) intolerance and children having secondary causes of malnutrition like cerebral palsy, congenital heart defects and cleft lip and palate were excluded from the study.

### Sampling procedure and sample size

We exhaustively included all 6-59 months old children hospitalized to the SC from January 01, 2023 to July 31, 2023 and our final sample was 168 children.

### Variables of the study

#### Outcome variable

Recovery time

Event: Stabilized and transferred to out-patient therapeutic programme (OTP)

Censored: Death, left against medical advice (LAMA), and medical referrals

### Independent variables

- Main exposure variable: Type of SAM
- Other independent variables: Socio-demographic characteristics, routine medications, supplements and co-morbidities during admission

### Operational definitions

- **Recovery time or length of stay (LoS)**: refers to the number of days it takes from admission until the child got recovered from medical complications.
- **Stabilized and transferred to OTP**: 6-59 months old children can continue the nutritional rehabilitation phase at OTP once they become free of medical problems, grade three oedema and regained appetite.

- **Defaulted or left against medical advice (LAMA)**: Patients who are not found in SC for two consecutive days, or who left the hospital contrary to medical advice offered while the child has not recovered.
- **Death**: child who deceases while taking management in stabilization center.

### Data collection tools and data quality control

#### Data collection tool

Structured questionnaire was generated referring to the FMoH SAM management protocol. The questionnaire was attached with the patient file and needed information was collected as stated in the important documents like SAM registration logbook, SAM monitoring multi-chart and patient clinical files.

#### Data quality assurance

The structured questionnaire was prepared in English, same language used by the Ethiopian SAM treatment protocol. It is pretested in 17 patients from a different health facility (ten per cent of our total sample), was reviewed for sequence and layout. After receiving one-day training on data collection, the nurses, nutrition nurses and nutrition supervisor were in charge of completing the questionnaire along with their routine activities. The collected data was checked by the nutrition supervisor for its accuracy, completeness & consistency and remedial actions were taken on spot.

#### Data processing and analysis

The collected data got coded and entered into Microsoft Excel spreadsheet 2016 version and then exported to statistical package for social sciences (SPSS) version 25 for analysis. For analytical convenience, variables were transformed and recoded into numeric variables. Data was cleaned earlier to analysis by exploring levels of missing values and applied standardized Z score to check presence of significant outliers and did not found any extreme values. Variance inflation factor (VIF) and levels of tolerance was used to assess multicollinearity among independent variables and none of them was suggestive of significant correlation.

Descriptive analysis is executed and presented in tables and percentages along with the appropriate central tendency measure (median survival time in days). Kaplan-Meier & Cox regression was applied to determine the association of independent variables with dependent. Length of stay was (LoS) estimated using Kaplan-Meier procedure with Log Rank (Mantel-Cox) test to examine whether the observed difference of LoS between different groups of predictor variables is significant or not. Chi-square test was done to determine if there were adequate cell counts for each categorical variable. Independent variables with *p-value* of <0.2 during the bivariate Cox regression were nominated for multivariable Cox regression.

The proportional hazard assumption was checked by examining plots of LoS for model variables. The plotted points approximately lie nearby a line that has unit slope and zero intercept. Overall fitness of the model was assured by Omnibus tests of model coefficients at 5% level of significance. With entry and removal probabilities of 0.05 and 0.1 respectively, multivariable Cox regression was run using Stepwise Backward Wald method to detect independent predictors of recovery time. Finally, adjusted hazard ratio (AHR) with 95% confidence interval (CI) was used to show the strength of association and declare a statistical significance at p-value of < 0.05.

## Results

From the total 194 children aged 6-59 months admitted to suhul hospital with complicated SAM, 168 were enrolled in this study while the remaining 26 were ineligible due to the exclusion criteria. Recovery rate was 94% while the rest six per cent of the participants were censored.

### Socio-demographic characteristics and baseline clinical conditions at admission

With mean and median age of 21.65 and 18 months respectively, majority (68.5%) of the participants fall under the 6-24 months old category while 54.2 per cent out of the whole cohort were females. About 29.2 per cent of patients were from IDPs and most of participants were new admissions (97%). Ninety-six children (57.1 per cent) of children were breastfed while one fourth of them had edematous type of SAM at admission. Dehydration, failed appetite, pneumonia, persistent diarrhea, hyperthermia, malaria and skin lesions found to be main contributors of hospitalization whereas anemia, unconsciousness and vomiting were less encountered complications comparatively. (**Table 1)**

**Table 1:**
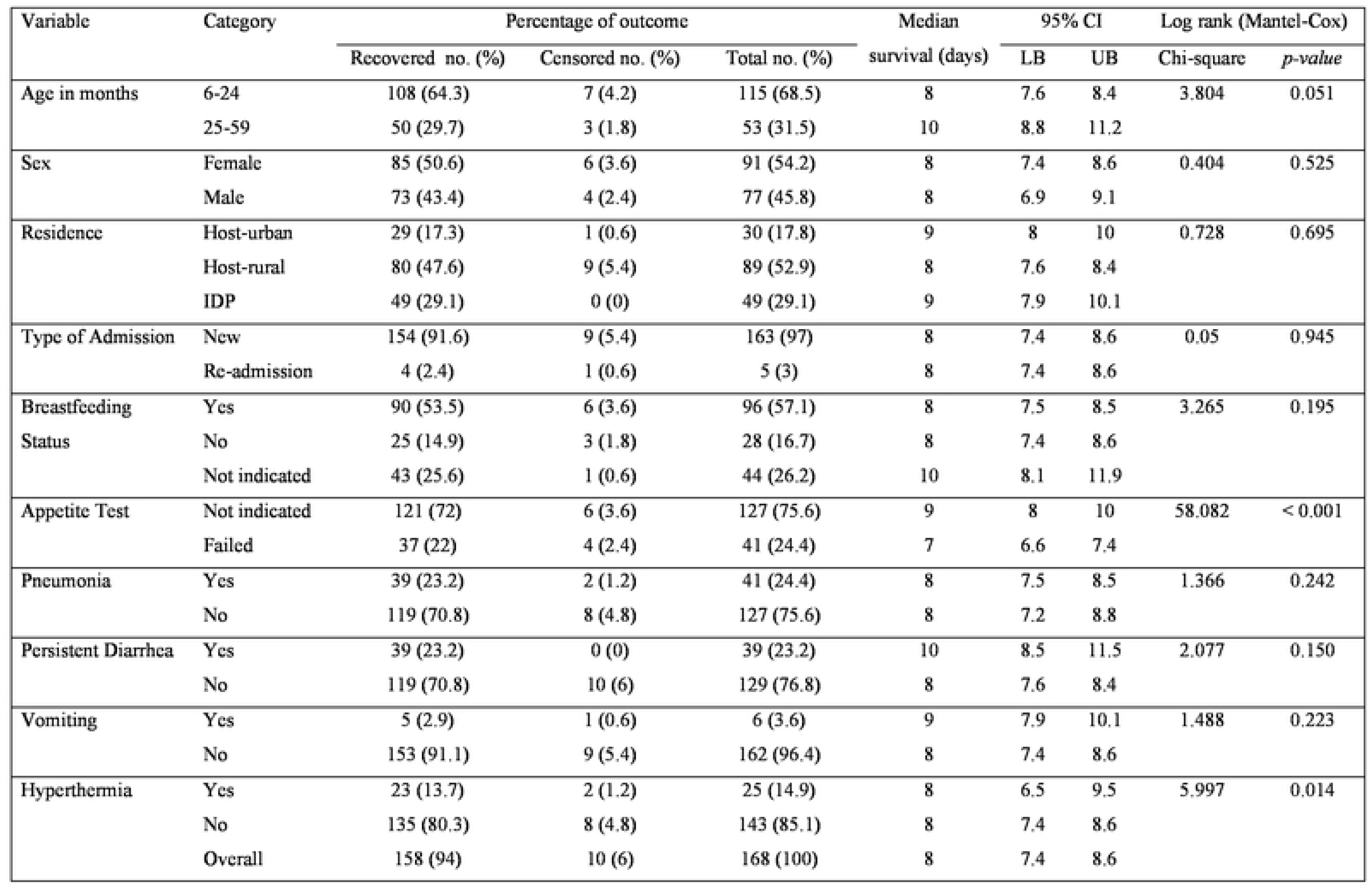
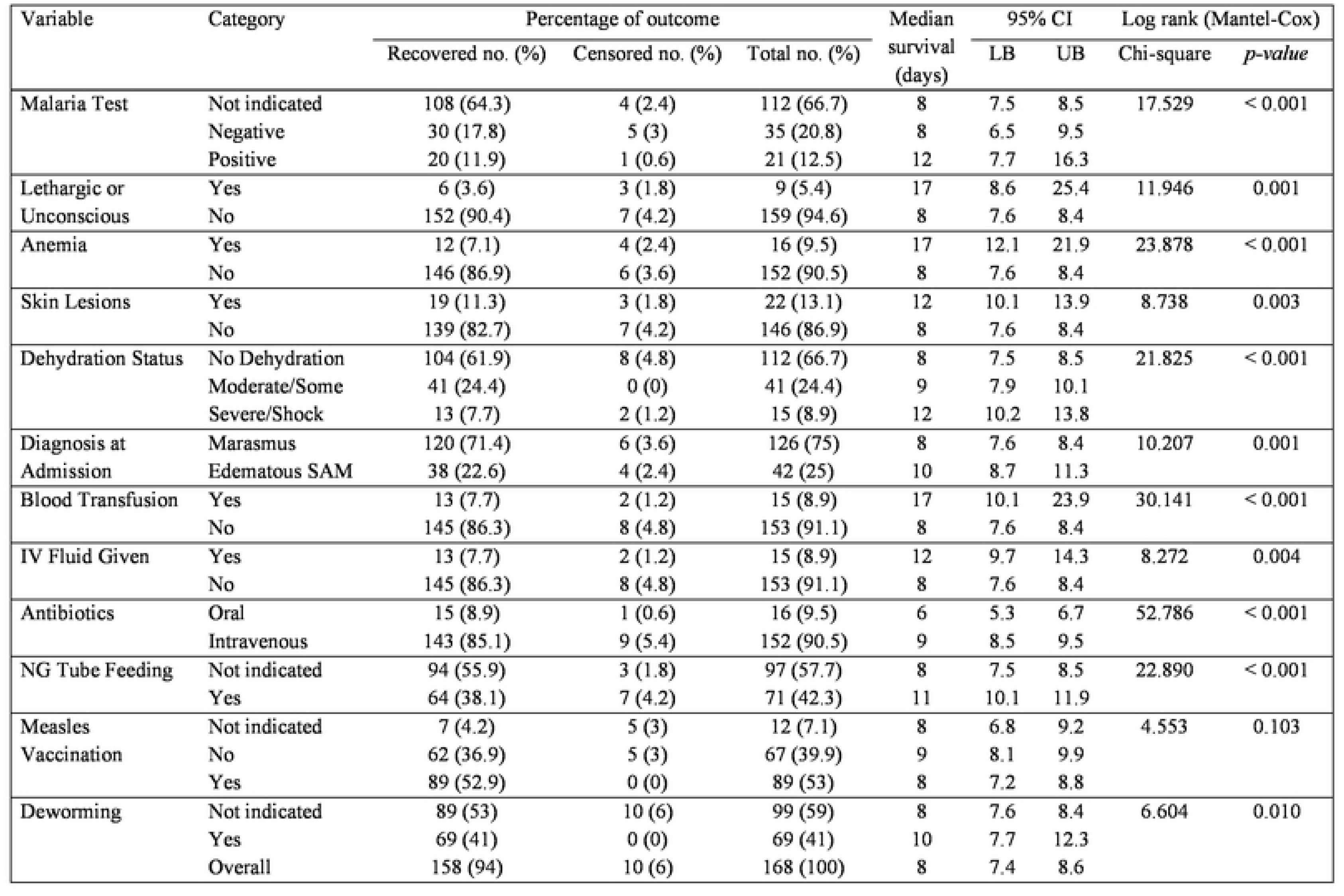
Log Rank (Mantel-Cox) test of equality of survival distributions for sociodemographic characteristics, clinical conditions at admission, routine medications and feeding in chidrcn with severe undernutrition treated in suhul hospital, 2023.

| Variable | Category | Percentage of outcome |  |  | Median survival (days) | 95% CI |  | Log rank (Mantel-Cox) |  |
| --- | --- | --- | --- | --- | --- | --- | --- | --- | --- |
|  |  | Recovered no. (%) | Censored no. (%) | Total no. (%) |  | LB | UB | Chi-square | *p-value* |
| Age in months | 6-24 | 108 (64.3) | 7 (4.2) | 115 (68.5) | 8 | 7.6 | 8.4 | 3.804 | 0.051 |
|  | 25-59 | 50 (29.7) | 3 (1.8) | 53 (31.5) | 10 | 8.8 | 11.2 |  |  |
| Sex | Female | 85 (50.6) | 6 (3.6) | 91 (54.2) | 8 | 7.4 | 8.6 | 0.404 | 0.525 |
|  | Male | 73 (43.4) | 4 (2.4) | 77 (45.8) | 8 | 6.9 | 9.1 |  |  |
| Residence | Host-urban | 29 (17.3) | 1 (0.6) | 30 (17.8) | 9 | 8 | 10 | 0.728 | 0.695 |
|  | Host-rural | 80 (47.6) | 9 (5.4) | 89 (52.9) | 8 | 7.6 | 8.4 |  |  |
|  | IDP | 49 (29.1) | 0 (0) | 49 (29.1) | 9 | 7.9 | 10.1 |  |  |
| Type of Admission | New | 154 (91.6) | 9 (5.4) | 163 (97) | 8 | 7.4 | 8.6 | 0.05 | 0.945 |
|  | Re-admission | 4 (2.4) | 1 (0.6) | 5 (3) | 8 | 7.4 | 8.6 |  |  |
| Breastfeeding Status | Yes | 90 (53.5) | 6 (3.6) | 96 (57.1) | 8 | 7.5 | 8.5 | 3.265 | 0.195 |
|  | No | 25 (14.9) | 3 (1.8) | 28 (16.7) | 8 | 7.4 | 8.6 |  |  |
|  | Not indicated | 43 (25.6) | 1 (0.6) | 44 (26.2) | 10 | 8.1 | 11.9 |  |  |
| Appetite Test | Not indicated | 121 (72) | 6 (3.6) | 127 (75.6) | 9 | 8 | 10 | 58.082 | < 0.001 |
|  | Failed | 37 (22) | 4 (2.4) | 41 (24.4) | 7 | 6.6 | 7.4 |  |  |
| Pneumonia | Yes | 39 (23.2) | 2 (1.2) | 41 (24.4) | 8 | 7.5 | 8.5 | 1.366 | 0.242 |
|  | No | 119 (70.8) | 8 (4.8) | 127 (75.6) | 8 | 7.2 | 8.8 |  |  |
| Persistent Diarrhea | Yes | 39 (23.2) | 0 (0) | 39 (23.2) | 10 | 8.5 | 11.5 | 2.077 | 0.150 |
|  | No | 119 (70.8) | 10 (6) | 129 (76.8) | 8 | 7.6 | 8.4 |  |  |
| Vomiting | Yes | 5 (2.9) | 1 (0.6) | 6 (3.6) | 9 | 7.9 | 10.1 | 1.488 | 0.223 |
|  | No | 153 (91.1) | 9 (5.4) | 162 (96.4) | 8 | 7.4 | 8.6 |  |  |
| Hyperthermia | Yes | 23 (13.7) | 2 (1.2) | 25 (14.9) | 8 | 6.5 | 9.5 | 5.997 | 0.014 |
|  | No | 135 (80.3) | 8 (4.8) | 143 (85.1) | 8 | 7.4 | 8.6 |  |  |
|  | Overall | 158 (94) | 10 (6) | 168 (100) | 8 | 7.4 | 8.6 |  |  |
|  |  | Recovered no. (%) | Censored no. (%) | Total no. (%) |  | LB | UB | Chi-square | *p-value* |
| Malaria Test | Not indicated | 108 (64.3) | 4 (2.4) | 112 (66.7) | 8 | 7.5 | 8.5 | 17.529 | < 0.001 |
|  | Negative | 30 (17.8) | 5 (3) | 35 (20.8) | 8 | 6.5 | 9.5 |  |  |
|  | Positive | 20 (11.9) | 1 (0.6) | 21 (12.5) | 12 | 7.7 | 16.3 |  |  |
| Lethargic or Unconscious | Yes | 6 (3.6) | 3 (1.8) | 9 (5.4) | 17 | 8.6 | 25.4 | 11.946 | 0.001 |
|  | No | 152 (90.4) | 7 (4.2) | 159 (94.6) | 8 | 7.6 | 8.4 |  |  |
| Anemia | Yes | 12 (7.1) | 4 (2.4) | 16 (9.5) | 17 | 12.1 | 21.9 | 23.878 | < 0.001 |
|  | No | 146 (86.9) | 6 (3.6) | 152 (90.5) | 8 | 7.6 | 8.4 |  |  |
| Skin Lesions | Yes | 19 (11.3) | 3 (1.8) | 22 (13.1) | 12 | 10.1 | 13.9 | 8.738 | 0.003 |
|  | No | 139 (82.7) | 7 (4.2) | 146 (86.9) | 8 | 7.6 | 8.4 |  |  |
| Dehydration Status | No Dehydration | 104 (61.9) | 8 (4.8) | 112 (66.7) | 8 | 7.5 | 8.5 | 21.825 | < 0.001 |
|  | Moderate/Some | 41 (24.4) | 0 (0) | 41 (24.4) | 9 | 7.9 | 10.1 |  |  |
|  | Severe/Shock | 13 (7.7) | 2 (1.2) | 15 (8.9) | 12 | 10.2 | 13.8 |  |  |
| Diagnosis at Admission | Marasmus | 120 (71.4) | 6 (3.6) | 126 (75) | 8 | 7.6 | 8.4 | 10.207 | 0.001 |
|  | Edematous SAM | 38 (22.6) | 4 (2.4) | 42 (25) | 10 | 8.7 | 11.3 |  |  |
| Blood Transfusion | Yes | 13 (7.7) | 2 (1.2) | 15 (8.9) | 17 | 10.1 | 23.9 | 30.141 | < 0.001 |
|  | No | 145 (86.3) | 8 (4.8) | 153 (91.1) | 8 | 7.6 | 8.4 |  |  |
| IV Fluid Given | Yes | 13 (7.7) | 2 (1.2) | 15 (8.9) | 12 | 9.7 | 14.3 | 8.272 | 0.004 |
|  | No | 145 (86.3) | 8 (4.8) | 153 (91.1) | 8 | 7.6 | 8.4 |  |  |
| Antibiotics | Oral | 15 (8.9) | 1 (0.6) | 16 (9.5) | 6 | 5.3 | 6.7 | 52.786 | < 0.001 |
|  | Intravenous | 143 (85.1) | 9 (5.4) | 152 (90.5) | 9 | 8.5 | 9.5 |  |  |
| NG Tube Feeding | Not indicated | 94 (55.9) | 3 (1.8) | 97 (57.7) | 8 | 7.5 | 8.5 | 22.890 | < 0.001 |
|  | Yes | 64 (38.1) | 7 (4.2) | 71 (42.3) | 11 | 10.1 | 11.9 |  |  |
| Measles Vaccination | Not indicated | 7 (4.2) | 5 (3) | 12 (7.1) | 8 | 6.8 | 9.2 | 4.553 | 0.103 |
|  | No | 62 (36.9) | 5 (3) | 67 (39.9) | 9 | 8.1 | 9.9 |  |  |
|  | Yes | 89 (52.9) | 0 (0) | 89 (53) | 8 | 7.2 | 8.8 |  |  |
| Deworming | Not indicated | 89 (53) | 10 (6) | 99 (59) | 8 | 7.6 | 8.4 | 6.604 | 0.010 |
|  | Yes | 69 (41) | 0 (0) | 69 (41) | 10 | 7.7 | 12.3 |  |  |
|  | Overall | 158 (94) | 10 (6) | 168 (100) | 8 | 7.4 | 8.6 |  |  |

### Treatment, feeding and final outcome

Fifteen patients received blood transfusion, exactly the same number of patients were also resuscitated with intravenous (IV) fluids. Only sixteen patients took oral antibiotics while the rest received their medications through IV line. Nasogastric (NG) tube was inserted to administer therapeutic milk and rehydration solutions in seventy-one patients. Out of the 156 children eligible for measles vaccination, only 89 of them got vaccinated because of lack of the antigen. However, all children in the appropriate age for deworming (≥ 24 months) took single dose of albendazole or mebendazole. 95.2% of patients hospitalized with severe wasting were recovered whereas the recovery rate for children having edematous type of SAM at admission was 90.5%. Finally, 158 patients (94%) got stabilized and transferred to OTP whereas four (2.4%) and six (3.6%) were defaulted and dead respectively. The overall median length of stay until final outcome was 8 days (95% CI: 7.4, 8.6). (**Figure 1**)

**Figure 1:**
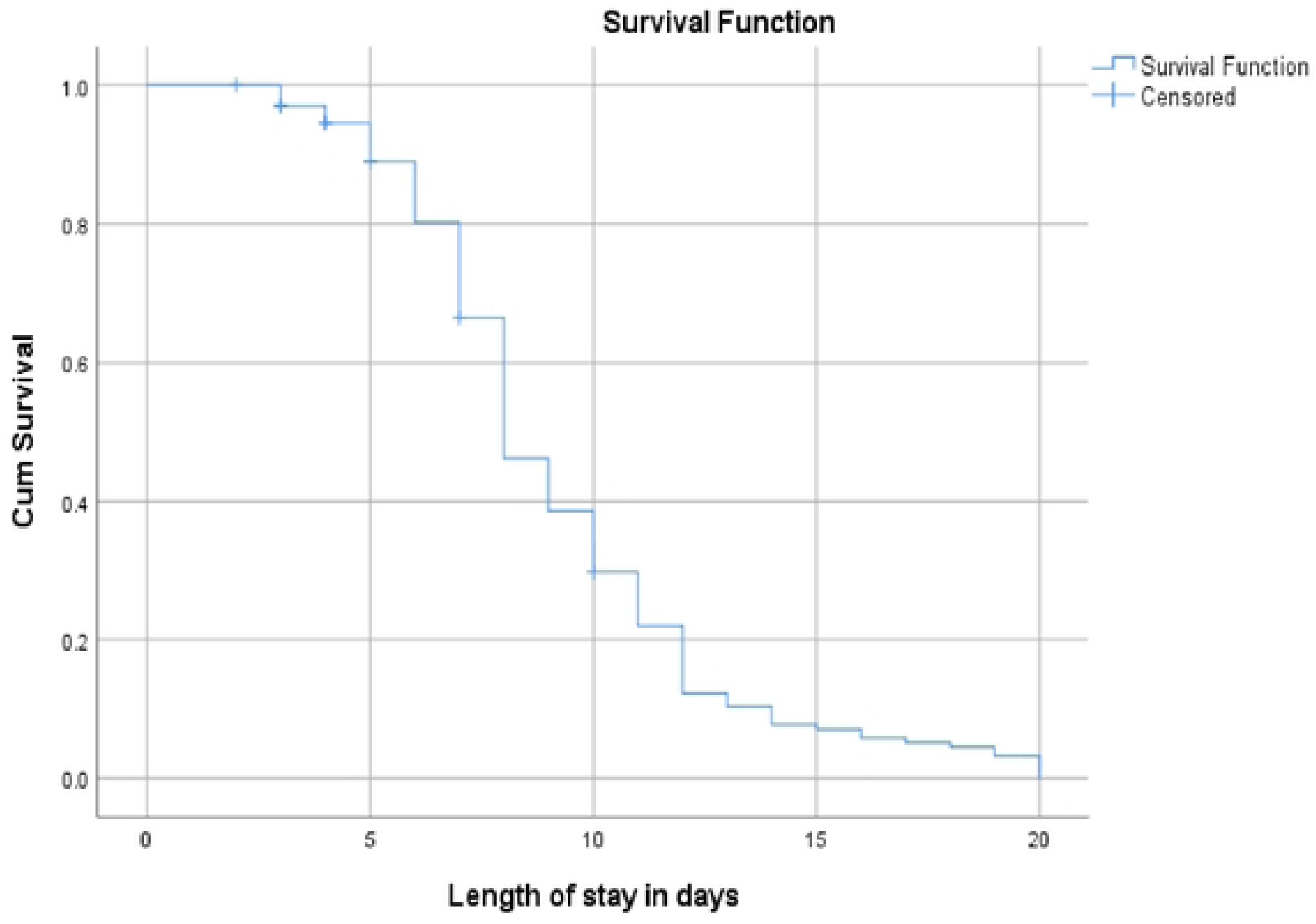
Final Kaplan Meier survival estimates of recovery time among 6-59 months old children treated for SAM in Suhul general hospital, 2023

### Kaplan Meier estimates of recovery time

Recovery time was estimated using Log Rank (Mantel-Cox) test of equality of survival distribution for all independent variables. (**Table 1**). The minimum, maximum, mean and median length of stay estimated to be two, twenty, eight point nine and eight days respectively. The LoS for children aged 25-59 months was higher than those aged 6-24 months. Nonetheless, this detected difference is by chance as it is not significant statistically (*p-value* 0.051). Similarly, we found some differences in LoS in residence of patient, breastfeeding status and persistent diarrhea with no statistical significance. However, the observed LoS differences in variables like type of SAM at admission (**Figure 2**), appetite test (**Figure 3**), hyperthermia, malaria status, anemia, blood transfusion (**Figure 4**), type of antibiotics, dehydration (**Figure 5**), NG tube feeding, skin lesions and deworming were statistically significant. The recovery time for those having edematous type of SAM was 10 days (95% CI: 8.7, 11.3), significantly longer compared with children having marasmus. (**Figure 2**)

**Figure 2:**
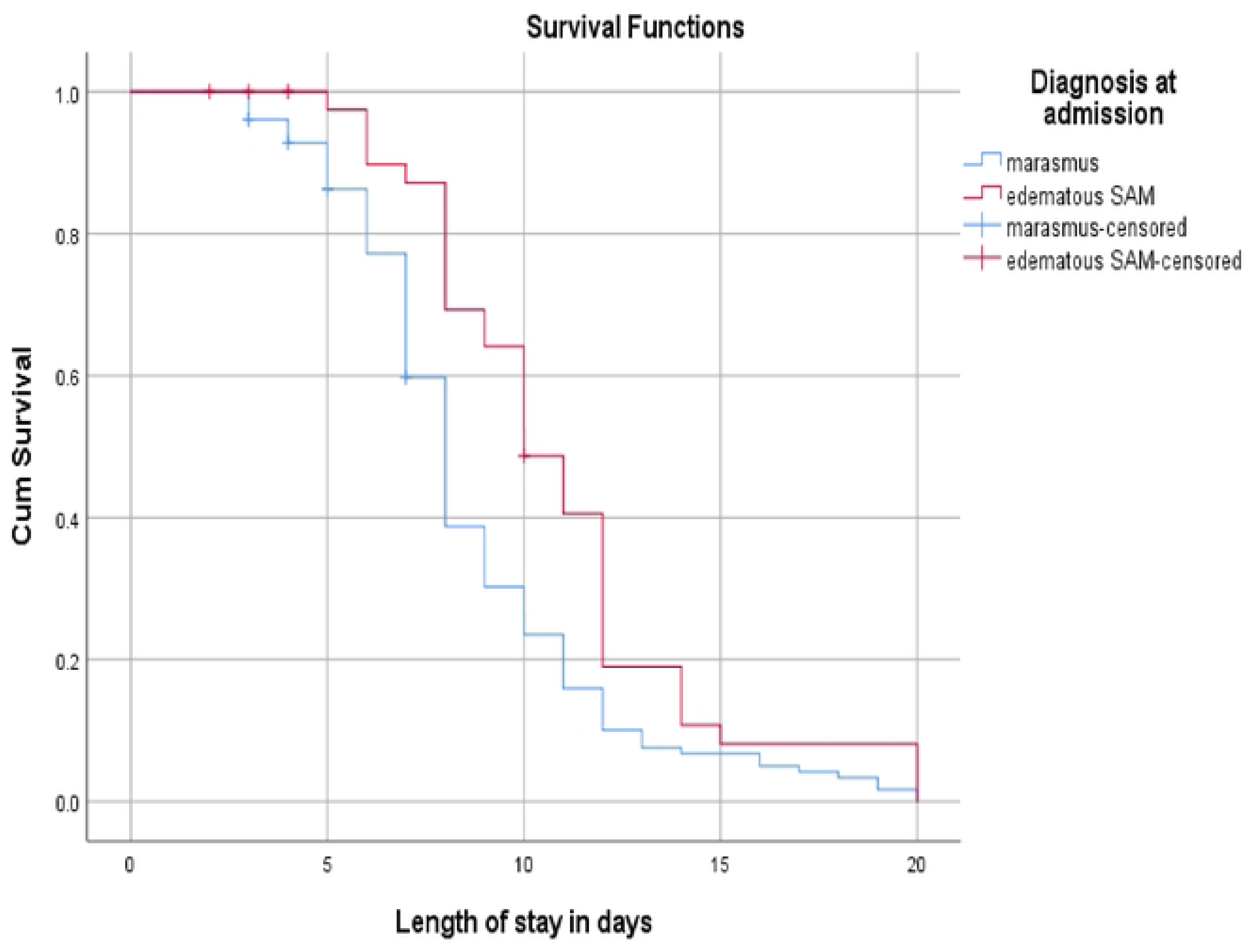
Kaplan Meier Survival curves of recovery time among 6-59 months old children by their diagnosis at admission, Suhul general hospital, 2023

**Figure 3:**
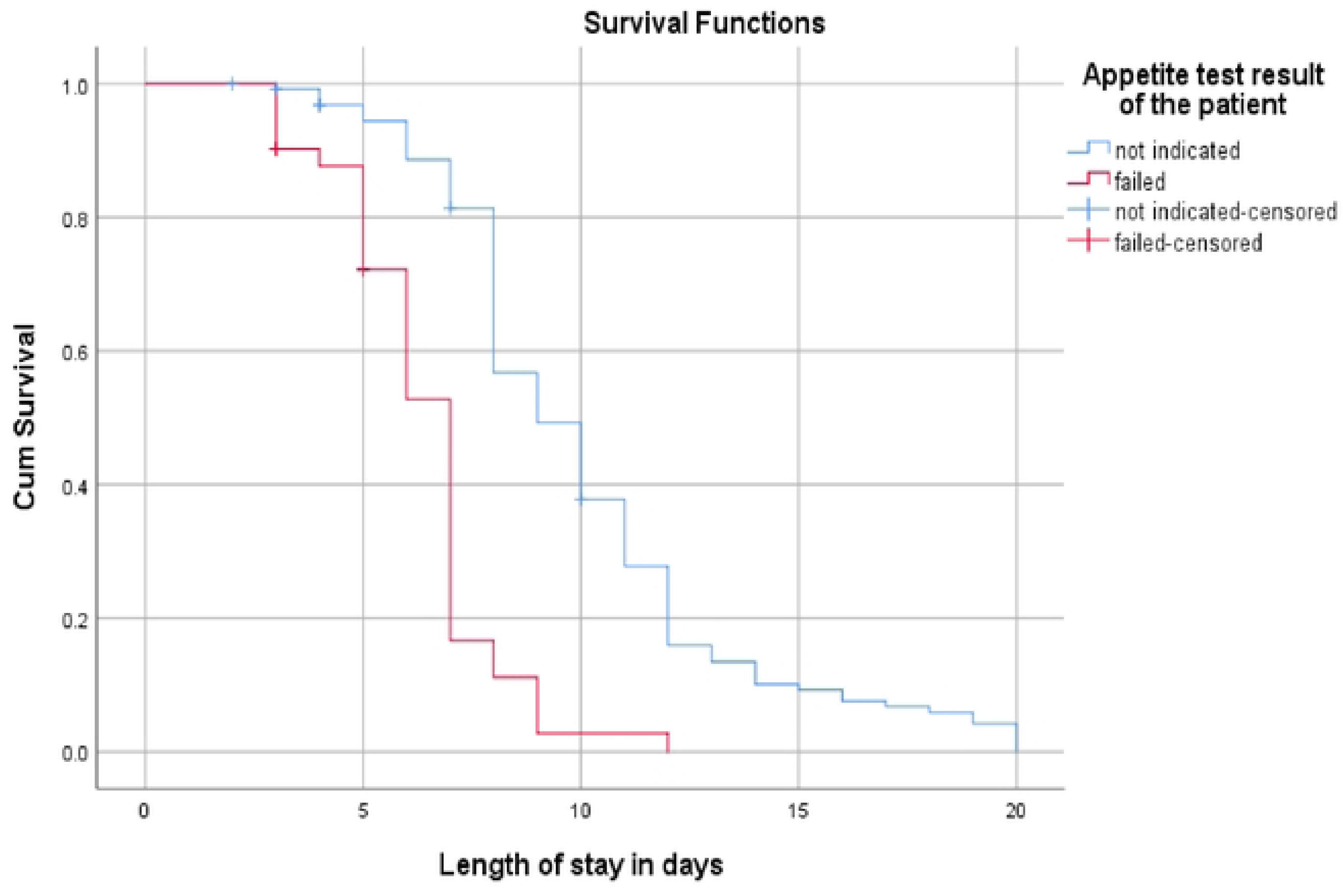
Kaplan Meier Survival estimates of recovery time among 6-59 months old children by their appetite test result at admission, Suhul general hospital, 2023

**Figure 4:**
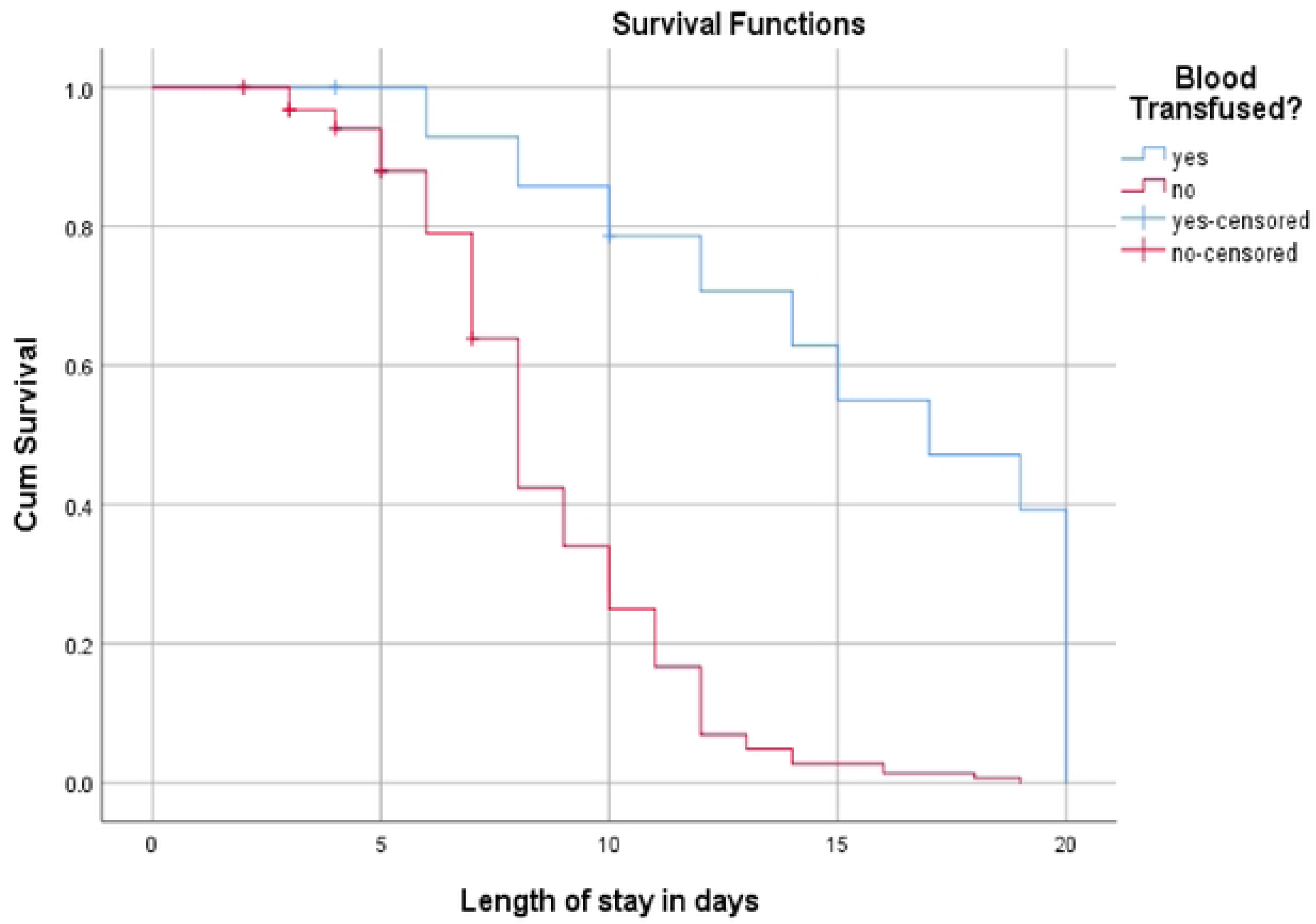
Kaplan Meier Survival curves of recovery time among 6-59 months old children based on blood transfusion after admission, Suhul general hospital, 2023

**Figure 5:**
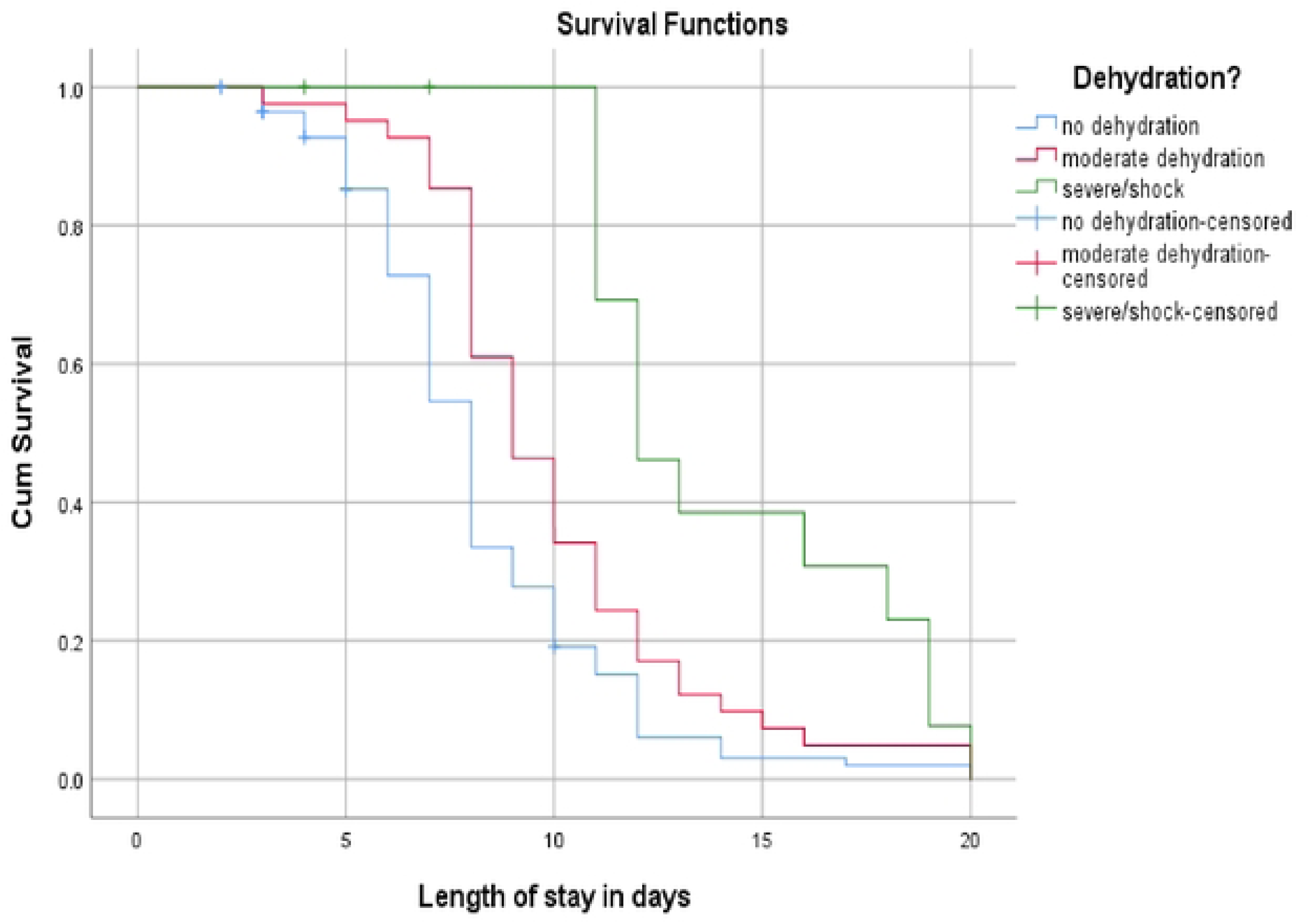
Kaplan Meier Survival curves of recovery time among 6-59 months old children by their level of at dehydration during admission, Suhul general hospital, 2023

### Predictors of recovery time

Fourteen independent variables with *p-value* < 0.2 during the bivariate Cox regression were included in the final multivariable Cox regression. Once validity of the model assumptions ascertained and amendment for potential confounders, six significant predictors of recovery time were identified, namely; appetite test, level of dehydration, diagnosis at admission, blood transfusion, antibiotics and NG tube feeding.

Patients hospitalized with failed appetite found to have about two times highly likely to stabilize compared to those admitted with other medical complications, for whom appetite test is not indicated (AHR = 1.874; 95% CI: 1.180-2.978). Level of dehydration resulted in 46.2% (AHR = 0.538; 95% CI: 0.361-0.800) and 75% (AHR = 0.250; 95% CI: 0.128-0.489) less likelihood of recovering from SAM in children being dehydrated and shocked at admission respectively. Compared with patients having severe wasting, those suffering from edematous SAM were less likely to recover by 55% (AHR = 0.452; 95% CI: 0.294-0.694). Patients who did not receive blood transfusion had more than fivefold likelihood of quicker recovery compared with their blood transfused counter parts (AHR = 5.559; 95% CI: 2.419-12.773).

Similarly, children treated with IV antibiotics and those who took the therapeutic milk via NG tube were 63.5 (AHR = 0.365; 95% CI: 0.192-0.692) and 46.9 per cent (AHR = 0.531; 95% CI: 0.372-0.758) less likely in recovering early compared to their counterparts who took oral antibiotics and who were able to take the therapeutic milk orally respectively. (**Table 2**)

**Table 2:** Predictors of recovery time among children treated for SAM in Suhul general hospital from January 01, 2023 to July 31, 2023.

| Variable | Category | Percentage of outcome |  |  | CHR (95% CI) | AHR (95% CI) | *P-value* |
| --- | --- | --- | --- | --- | --- | --- | --- |
|  |  | Recovered | Censored | Total |  |  |  |
| Age in months | 6-24 | 108 | 7 | 115 | 1 | 1 | 0.591 |
|  | 25-59 | 50 | 3 | 53 | 0.745 (0.531-1.045) | 0.900 (0.612-1.322) |  |
| Appetite test | Not indicated | 121 | 6 | 127 | 1 | 1 | **0.008** |
|  | Failed | 37 | 4 | 41 | 3.934 (2.619-5.911) | 1.874 (1.180-2.978) |  |
| Hyperthermia | Yes | 23 | 2 | 25 | 1 | 1 | 0.948 |
|  | No | 135 | 8 | 143 | 1.670 (1.040-2.681) | 1.020 (0.565-1.839) |  |
| Malaria Test | Not indicated | 108 | 4 | 112 | 1 | 1 | 0.263 |
|  | Negative | 30 | 5 | 35 | 0.790 (0.526-1.186) | 1.045 (0.651-1.677) |  |
|  | Positive | 20 | 1 | 21 | 0.358 (0.204-0.629) | 0.430 (0.140-1.323) |  |
| Lethargy/unconsciousness | Yes | 6 | 3 | 9 | 1 | 1 | 0.512 |
|  | No | 152 | 7 | 159 | 3.271 (1.419-7.542) | 1.429 (0.429-4.149) |  |
| Anemia | Yes | 12 | 4 | 16 | 1 | 1 | 0.545 |
|  | No | 146 | 6 | 152 | 3.775 (1.992-7.153) | 0.546 (0.077-3.876) |  |
| Skin lesions | Yes | 19 | 3 | 22 | 1 | 1 | 0.370 |
|  | No | 139 | 7 | 146 | 1.862 (1.146-3.024) | 1.340 (0.707-2.540) |  |
| Level of dehydration | No dehydration | 104 | 8 | 112 | 1 | 1 | **< 0.001** |
|  | Moderate/Some | 41 | 0 | 41 | 0.650 (0.452-0.935) | 0.538 (0.361-0.800) |  |
|  | Severe/shock | 13 | 2 | 15 | 0.339 (0.188-0.611) | 0.250 (0.128-0.489) |  |
| Diagnosis at admission | Marasmus | 120 | 6 | 126 | 1 | 1 | **< 0.001** |
|  | Edematous SAM | 38 | 4 | 42 | 0.594 (0.411-0.860) | 0.452 (0.294-0.694) |  |
| Blood transfusion | Yes | 13 | 2 | 15 | 1 | 1 | **< 0.001** |
|  | No | 145 | 8 | 153 | 5.667 (2.652-12.110) | 5.559 (2.419-12.773) |  |
| IV fluid | Yes | 13 | 2 | 15 | 1 | 1 | 0.204 |
|  | No | 145 | 8 | 153 | 2.050 (1.154-3.643) | 1.785 (0.731-4.363) |  |
| Antibiotics | Oral | 15 | 1 | 16 | 1 | 1 | **0.002** |
|  | Intravenous (IV) | 143 | 9 | 152 | 0.162 (0.090-0.291) | 0.365 (0.192-0.692) |  |
| NG tube feeding | Not indicated | 94 | 3 | 97 | 1 | 1 | **0.001** |
|  | Yes | 64 | 7 | 71 | 0.503 (0.364-0.697) | 0.531 (0.372-0.758) |  |
| Deworming | Not indicated | 89 | 10 | 99 | 1 | 1 | 0.805 |
|  | Yes | 69 | 0 | 69 | 0.691 (0.500-0.954) | 1.076 (0.601-1.925) |  |

## Discussion

This research has assessed whether there is significant difference in recovery among different groups of patients and the overall recovery time from complicated SAM with its predictors. We have compared the reproducibility of our current findings with different studies of similar titles.

Hence, marasmus was the main provisional diagnosis at admission, consistent with reports from recent studies in Aksum (17), Addis Ababa (25) and Southern Ethiopia (6). The median recovery time was 8 days (95% CI: 7.4, 8.6) which fulfils the requirement for maximum LoS in SC by WHO to be less than four weeks (26). This finding is similar with prospective study conclusions from Sidama regional state of Ethiopia (27). Conversely, this is much better (shorter) compared to findings from different public hospitals in the capital city of the country and different institutions in Oromia (25,28), at which children were staying for about 19 and 21 days to be stabilized respectively. Such variability could be due to differences in adhering to the national guideline strictly and lack of trainings on the protocol as well as supply shortage among hospitals.

The rates of recovery, defaulter and death from the whole cohort were 94, 2.4 and 3.6 per cent respectively. This satisfies the performance indicators for SC set by the sphere association (29) and is way better achievement in reference to recent study findings from Jimma university medical center (30), that found only a quarter of the participants recovered. This is probably attributed to the comprehensive medical supplies interconnected with training, health promotion and supplementary IYCF services Suhul hospital was receiving from several humanitarian organizations during the crisis. Nonetheless, our achievements are similar with reports from Katsina state of Nigeria (31), a project received similar assistance from humanitarian NGO, achieved 95.7% of recovery and 4.3% death rate with LoS till outcome of 6 days.

In the final multivariable analysis using Cox proportional hazard regression, failed appetite test was significant predictor of time to recover, that in line with study results from Adama hospital (32). Likewise, we detected edematous type of SAM at admission resulted in reduced chance of earlier recovery by 55% compared to those hospitalized with marasmus. This is compatible with findings from Benishangul Gumuz (20) who declared children admitted with severe wasting had 1.7 times higher chance of recovering early than those with edema, Amhara regional state of Ethiopia (33) who concluded hospitalized with edematous type of SAM resulted in 41% lower likelihood of recovery and researchers from Uganda (34) as well as Nigeria (31) stated that edematous form of SAM had extended LoS till recovery. But this is contradicting to reports from southern Ethiopia (27,35). This might be due to the added co-morbidities either forms of SAM, which opens areas of further studies to figure out reasons.

Our conclusion of special treatments like blood transfusion and provision of IV antimicrobials as independent predictors of LoS is on the same page with findings of researchers from Hawassa tertiary hospital, Southern Ethiopia (36) and other African Nations (34,37). By the same token, our finding on NG tube feeding resulted in significantly longer recovery time is backed by several researchers (38–40).

## Conclusion and Recommendations

Bottom line of the study shows the inpatient therapeutic feeding center has met the agreed requirements (indicators) for nutrition interventions during humanitarian crises, which is relatively superior achievement compared with majority of latest studies in the thematic area. Appetite test, level of dehydration, diagnosis at admission, blood transfusion, type of antibiotics and nasogastric tube feeding were confirmed predictors of time till recovery from SAM.

Therefore, the hospital leadership along with the regional health bureau and other humanitarian agencies should give emphasis on training clinical workforce directly involved in severely undernourished child management and care, so that the clinicians will become skillful to identify and properly manage the main complications of SAM like severe anemia, dehydration and shock. Furthermore, the blood bank unit and the laboratory of the hospital should be looked after to ensure safe blood transfusion for those who need it.

## Data Availability

All relevant data are within the manuscript and its Supporting Information files.

## Abbreviations

AHR: Adjusted Hazard Ratio
CHR: Crude Hazard Ratio
CI: Confidence Interval
IDP: Internally Displaced Person
IV: Intravenous
LoS: Length of Stay
NG: Naso-Gastric
NGO: Non-Governmental Organization
OTP: Outpatient Therapeutic Programme
SAM: Severe Acute Malnutrition
SC: Stabilization Center

## Declarations

### Ethical approval and consent to participate

Complying by the declaration of Helsinki for studies involving human beings, this research was commenced after receiving expedited ethics approval from Aksum university, college of health sciences, health research ethics review board (HRERB) with reference number: **<u>IRB 009/2023</u>**. This also fulfills the ethical standards set by the country (41). Permission was obtained from those in charge of the hospital’s stabilization center to collect relevant data items for this study. Approved by HRERB in Aksum university, college of health sciences, informed written consent was taken from each participant’s caretaker (legal guardian).

### Consent for publication

Not applicable

### Availability of data and materials

The records used to yield analytical outputs for this study are attached along with the manuscript as supplemental file **(S1: anonymized SC data. SAV).** Additionally, we have attached the steps we applied in the final multivariable Cox regression model **(S2).**

### Authors’ contributions

WT proposed and designed the study. Analysis, writing up the manuscript and its final approval is done by both authors.

## Acknowledgements

Our earnest appreciation goes to the healthcare workers of the hospital who never gave up and were tirelessly serving their society with almost no medical supplies and without salaries due to the war for nearly two years. We are warmly grateful for the nurses assigned in the paediatrics ward for their cooperation in collecting the data appropriately.

